# Inferring Gender from First Names: Comparing the Accuracy of Genderize, Gender API, and the gender R Package on Authors of Diverse Nationality

**DOI:** 10.1101/2024.01.30.24302027

**Authors:** Alexander D. VanHelene, Ishaani Khatri, C. Beau Hilton, Sanjay Mishra, Ece D. Gamsiz Uzun, Jeremy L. Warner

## Abstract

Meta-researchers commonly leverage tools that infer gender from first names, especially when studying gender disparities. However, tools vary in their accuracy, ease of use, and cost. The objective of this study was to compare the accuracy and cost of the commercial software Genderize and Gender API, and the open-source gender R package. Differences in binary gender prediction accuracy between the three services were evaluated. Gender prediction accuracy was tested on a multi-national dataset of 32,968 gender-labeled clinical trial authors. Additionally, two datasets from previous studies with 5779 and 6131 names, respectively, were re-evaluated with modern implementations of Genderize and Gender API. The gender inference accuracy of Genderize and Gender API were compared, both with and without supplying trialists’ country of origin in the API call. The accuracy of the gender R package was only evaluated without supplying countries of origin since. The accuracy of Genderize, Gender API, and the gender R package were defined as the percentage of correct gender predictions. Accuracy differences between methods were evaluated using McNemar’s test. Genderize and Gender API demonstrated overall 96.6% and 96.1% accuracy, respectively, when countries of origin were not supplied in the API calls. Genderize and Gender API achieved the highest accuracy when predicting the gender of German authors with accuracies greater than 98%. Genderize and Gender API were least accurate with South Korean, Chinese, Singaporean, and Taiwanese authors, demonstrating below 82% accuracy. The gender R package achieved below 86% accuracy on the full dataset. In the replication studies, Genderize and gender API demonstrated better performance than in the original publications. Our results indicate that Genderize and Gender API are highly accurate, except when evaluating South Korean, Chinese, Singaporean, and Taiwanese names. We also demonstrated that Genderize can provide similar accuracy to Gender API while being 4.85x less expensive.

**Author Summary:** Gender disparities in academia have prompted researchers to investigate gender gaps in professorship roles and publication authorship. Of particular concern are the gender gaps in cancer clinical trial authorship. Methodologies that evaluate gender disparities in academia often rely on tools that infer gender from first names. Tools that predict gender from first names are often used in methodologies that determine the gender ratios of academic departments or publishing authors in a discipline. However, researchers must choose between different gender predicting tools that vary in their accuracy, ease of use, and cost. We evaluated the binary gender prediction accuracy of Genderize, Gender API, and the gender R package on a gold-standard dataset of 32,968 clinical trialists from around the world. Genderize and Gender API cost money to use, while the gender R package is free and open source. We found that Genderize and Gender API were more accurate than the gender R package. In addition, Genderize is cheaper than Gender API, but is more sensitive to inconsistencies in name formatting and the presence of diacritical marks. Both Genderize and Gender API were most accurate with western names.

## Introduction

One of the most well-documented disparities in STEM is gender disparity [1,2]. This issue is especially notable in the cancer clinical trial domain, with underrepresentation of women in the leadership of pivotal trials documented as recently as within the last decade [3]. The study of gender disparity in scientific authorship often requires the determination of gender from very limited data, e.g., author forenames. Software [4–9] that infers gender from forenames could potentially enable researchers to automate gender prediction in large datasets. Commercial gender prediction services [10,11] such as Genderize and Gender API programmatically predict gender from first names. The gender R package [12] is an open-source alternative to these proprietary gender prediction tools.

Gender prediction software has demonstrated high accuracy when evaluating Western first names, but often falters when evaluating names from Asian cultures [13]. Further, the presence of diacritical marks and hyphens reportedly affects the accuracy of gender prediction in some tools [14]. Few studies [15] to date have evaluated differences in accuracy in gender predicting software between Western and non-Western names. To our knowledge, no studies have evaluated how different ways of delimiting two-part first names e.g. Jean-Pierre vs Jean Pierre vs Jeanpierre, affect gender prediction accuracy.

We compared the gender prediction accuracy of Genderize, Gender API, and the gender R package using a large manually curated registry of cancer clinical trialists with labeled genders and diverse nationalities. In addition, we quantified the accuracy of these tools by author nationality and compared different strategies for delimiting two-part forenames, which are common in the English language spelling of Korean, Chinese, Singaporean, and Taiwanese names.

## Materials and Methods

Three gender prediction tools: 1) Genderize; 2) Gender API; and 3) the gender R package, were tested on a gold-standard registry of cancer clinical trialists with manually determined binary gender. Trialists’ names and affiliations were sourced from the HemOnc knowledge base, [16] a continually growing resource created to capture the standard-of-care treatments in the fields of hematology and oncology. The binary gender classifications used in our study refer to socially constructed gender categories, not biological sex [17,18]. Names in HemOnc are primarily sourced from the MEDLINE records of published clinical trials and undergo extensive normalization to account for the presence of diacritics, middle initials, misspellings, multipart last names represented as middle names, and other variations. When first names are not available through MEDLINE, the original manuscripts are examined for this information. Binary gender is determined by a combination of automated mappings of typically masculine or feminine forenames (e.g., John; Rebecca), web searches of publicly available information such as biographies on academic web pages, and consensus determinations including consultation with native speakers. If gender cannot be determined after these efforts, the author is labeled as “unknown gender”. A subset of journals does not provide forenames; in these cases, the gender is labeled as “could not be determined.” Country affiliations sourced from MEDLINE also undergo extensive normalization.

Gender prediction accuracy was defined as the percent of individuals whose gender was correctly predicted, as compared to the gold standard dataset. The percent of incorrect gender predictions and the percent of names with no predicted gender were also calculated. For binary statistical tests, gender predictions were categorized as successes or failures – correct gender predictions were defined as successes, while names with incorrect or absent predictions were failures.

All trialists with a gender determination were evaluated with Genderize and Gender API on 2023-11-21 using the R package httr (version 1.4.7). Both US Social Security Administration (SSA) and US Census Integrated Public Use Microdata Series (IPUMS) name datasets were used as a reference when predicting names with the gender R package [12] (version 0.6.0).

Genderize and Gender API were used to predict names with and without supplying a country of origin for the subset of authors with a singular country of affiliation. The Gender R package was only tested without supplying country names because the SSA and IPUMS methods do not provide that functionality. Two-part names were concatenated without any delimiter e.g. Jean-Pierre was converted to jeanpierre. Middle names were removed, unless an author had a first initial/middle name, in which case their middle name was used. Gender bias in name prediction was descriptively evaluated by calculating the percent of names that were misgendered, compared to the gold standard labeled dataset. In an additional analysis, accuracy differences resulting from delimiting two-part first names with different characters were evaluated. Two-part first name prediction accuracy was also evaluated using the first half of two-part names only. For example, the name Jean-Pierre was tested four ways: 1) jean-pierre; 2) jean pierre; 3) jeanpierre; and 4) jean.

In addition to predicting the gender of a first name, Genderize and Gender API also report an estimated probability that a gender prediction is correct. We evaluated the correlation between these API-reported probability estimates and the gold standard labeled dataset with linear regressions and Brier scores. Names with a reported probability less than or equal to 50% were excluded from the regression and Brier scores.

The gender prediction accuracies of Genderize and Gender API were also separately evaluated using publicly available datasets from two studies [15,19] that tested gender prediction in 2018 and 2021, respectively. The dataset [20] provided by Santamaria 2018 consisted of 5,779 names sourced from various other datasets. The dataset [21] sourced from Sebo 2021 consisted of 6,131 Swiss physicians. The names from these public datasets were not modified prior to our evaluation on 2023-11-07. Nor were nationalities supplied to Genderize and Gender API when evaluating these public datasets, following the original experimental design.

All software accuracy comparisons were computed in R version 4.3.1. Differences in accuracy between methods were evaluated using the default R stats package implementation of McNemar’s test [22]. Data analysis was facilitated with tidyverse [23] (version 2.0.0), haven [24] (version 2.5.3), readxl [25] (version 1.4.3), testthat [26] (version 3.1.10), ggpmisc [27] (version 0.5.5), and patchwork [28] (version 1.1.3) R libraries.

## Results

Out of 40,273 unique clinical trialists present in the HemOnc KB as of 2023-11-21, 37,420 (92.9%) had a resolvable first name and were thus eligible for gender determination. This group was sourced from 7,473 clinical trial manuscripts published between 1947-2023. After excluding trialists with gender not yet determined (n=4,360, 11.7%), those with a determined unknown gender (n=78, 0.2%), and those with a determined gender but initial-only first names (n=14, <0.1%), the final analysis set included 32,968 trialists with predetermined binary gender. Of the 32,968 trialists, 11,398 (34.6%) were designated as women. There were 7849 unique names after normalizing first initial/middle name combinations to only include a middle name. The remainder of names were shared by more than one individual. Michael was the most common name, with 473 (1.4%) occurrences. Only 1,899 (24.2%) of names occurred more than twice.

Of 25,240 trialists with a known site affiliation, 24,930 (98.8%) were affiliated with sites in a single country and were assigned to the country of their affiliated institution when querying Genderize and Gender API with nationalities. When excluding clinical trialists without a recorded country of origin, the number of trialists and unique names was 24,930 and 6,756, respectively. The final analysis set included trialists from 87 countries, the most abundant being the US with 9,485 (38%) affiliated trialists. There were 7,569 first name-country combinations that occurred only once. The most common first-name-country combination was David-US with 201 (0.8%) instances. Only 1,760 (7.1%) of first name-country combinations appeared more than twice. The 100 most common trialist name-country combinations are presented in **Supplementary Table 1**.

### Gender prediction accuracy when country of origin was not supplied (baseline case)

The overall accuracy of Genderize when predicting gender for the full dataset without supplying country was 96.6% with 2.3% incorrect gender predictions and 1.1% of names yielding no prediction (**Table 1**). Similarly, the overall accuracy of Gender API was 96.1% with 2.7% incorrect gender predictions and 1.1% of names resulting in no prediction. The accuracy of the gender R package’s predictions was lower, with 79.8% and 85.7% accuracy with the IPUMS and SSA methods, respectively. Names of men were misgendered as women less than 3% of the time for all gender prediction tools (**Table 1)**. Names of women were misgendered over 3% of the time for all services except the gender R package when using SSA data as a reference. The difference in the percent of correct gender predictions between Genderize and Gender API was significant in favor of Genderize (p<0.001). Likewise, the accuracy difference between the gender R package methods were also significant (p<0.001), in favor of the SSA method. Gender API demonstrated higher gender prediction accuracy when two-part names were delimited with a space: the percent of correctly inferred genders rose from 96.1% to 96.3%.

**Table 1:** Accuracy of Gender Predictions on 32,968 Included Trialists.

| Method<sup>a</sup> | Correct, n (%) | Incorrect, n (%) | No Predictions, n (%) | Men Incorrectly Gendered as Women, n (%)<sup>b</sup> | Women Incorrectly Gendered as Men, n (%)<sup>b</sup> |
| --- | --- | --- | --- | --- | --- |
| Genderize | 31,857/32,968 (96.6%) | 763/32,968 (2.3%) | 348/32,968 (1.1%) | 401/21,324 (1.9%) | 362/11,296 (3.2%) |
| Gender API | 31,690/32,968 (96.1%) | 899/32,968 (2.7%) | 379/32,968 (1.1%) | 393/21,320 (1.8%) | 506/11,269 (4.5%) |
| gender (IPUMS) | 26,294/32,968 (79.8%) | 1366/32,968 (4.1%) | 5308/32,968 (16.1%) | 508/17,941 (2.8%) | 858/9719 (8.8%) |
| gender (SSA) | 28,266/32,968 (85.7%) | 590/32,968 (1.8%) | 4112/32,968 (12.5%) | 489/18,595 (2.6%) | 101/10,261 (1%) |

<sup>a</sup>Two-part first names were appended together without a delimiting character.
<sup>b</sup>Denominators are not consistent across rows because names that did not return a gender prediction for a given service were excluded.

After restricting Genderize’s predictions to trialists affiliated with a single country, the percentage of correct, incorrect, and missing predictions were 96.2%, 2.6%, and 1.2% respectively (**Fig 1A**). Genderize achieved the highest accuracy when evaluating first names from German authors, and the lowest accuracy when evaluating names from South Korean, Chinese, Singaporean, and Taiwanese authors. When evaluating the same 24,929 clinical trialists with Gender API, the percentage of correct, incorrect, and missing predictions were 95.8%, 3%, and 1.3% respectively. Gender API also had high accuracy when predicting the gender of German authors, and the lowest accuracy when evaluating names from South Korean, Chinese, Singaporean, and Taiwanese authors. The difference in accuracy between Genderize and Gender API is significant (p<0.001), in favor of Genderize.

**Fig 1:**
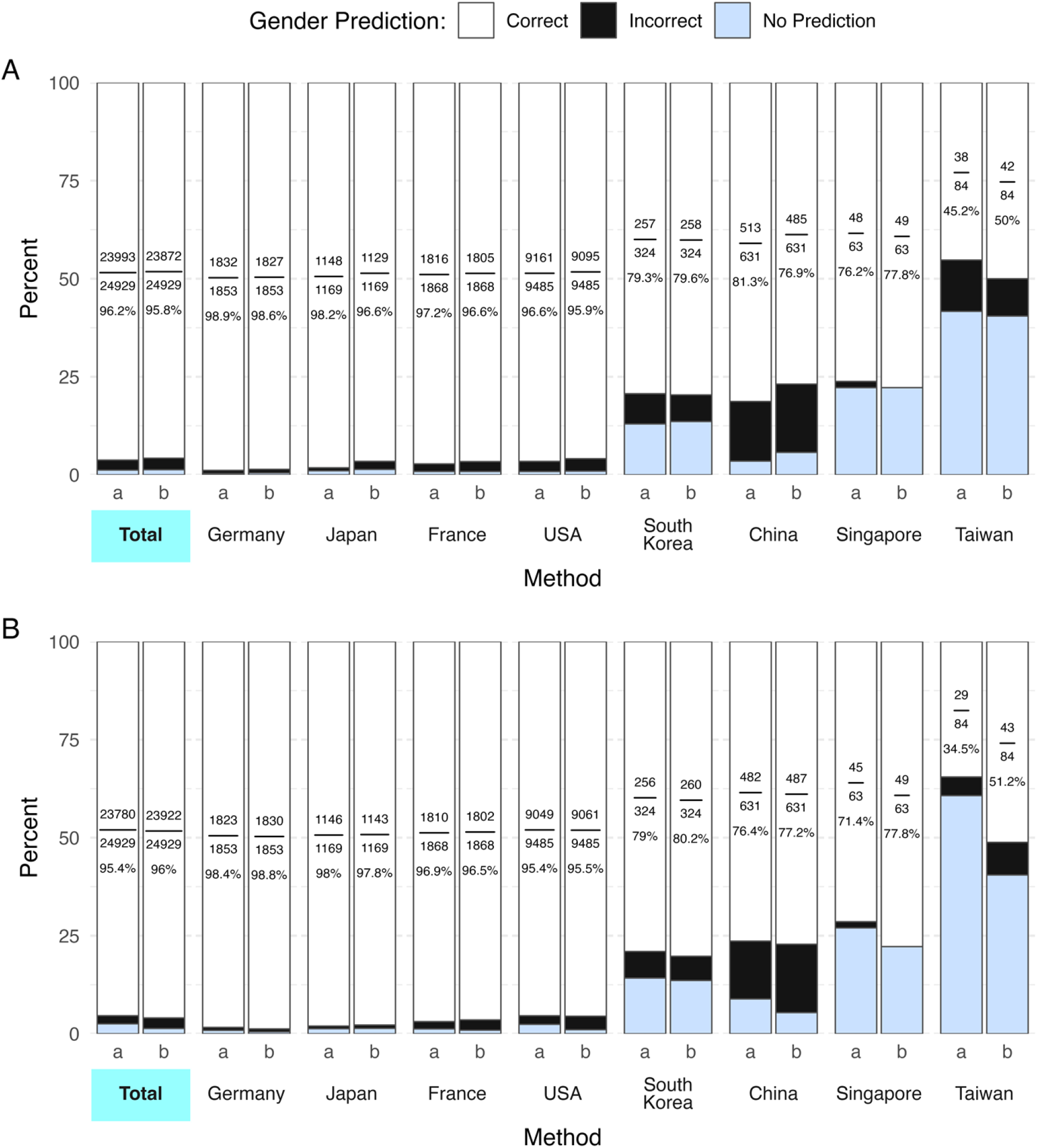
Accuracy of Gender Predictions. Panel A shows the gender prediction accuracies when countries are not included in the API call. Panel B shows the results when countries are included in the API call. The top 4 countries with the most trialists and all Asian countries are plotted. Method a is genderize and method b is Gender API. Each bar is labeled with the fraction and percentage of correct gender predictions. Two-part first names were appended together without a delimiting character.

### Gender prediction accuracy when country of origin was supplied to the API

The gender prediction accuracies when countries of origin were supplied to Genderize and Gender API are visualized in **Fig 1B**. Supplying the countries of origin alongside first names in the API call decreased the percentage of correct gender predictions when using Genderize from 96.2% to 95.4%, while also reducing the percentage of incorrect predictions from 2.6% to 2.1%. Conversely, including countries of origin increased the ratio of correct gender predictions of Gender API from 95.8% to 96% and decreased incorrect predictions from 3% to 2.7%. Supplying countries also increased the percentage of names with no gender prediction for Genderize from 1.2% to 2.5%, while Gender API remained constant at 1.3%. The difference in accuracy between Genderize and Gender API was significant in favor of Gender API (p<0.001).

### Gender Prediction accuracy when using different characters to delimit two-part forenames

Gender prediction accuracy when evaluating two-part names was higher when countries were not included in the API call in all contexts except when calling Genderize with the first half of a two-part name, e.g., Jean-Pierre as jean. Genderize was most accurate (76.4%) when no character was used to delimit two-part names, e.g., Jean-Pierre represented as jeanpierre (**Fig 2**). Genderize provided zero predictions for two-part first names delimited with a space. In contrast, Gender API achieved the highest gender prediction accuracy when delimiting two-part names with a space (83.5%). Gender prediction accuracy for two-part names was worse than for one-part names when countries were not included in the API call and two-part names were separated without a delimiter: OR 0.07 (95% CI 0.06-0.08) for Genderize and OR 0.08 (95% CI 0.07-0.09) for Gender API, respectively.

**Fig 2:**
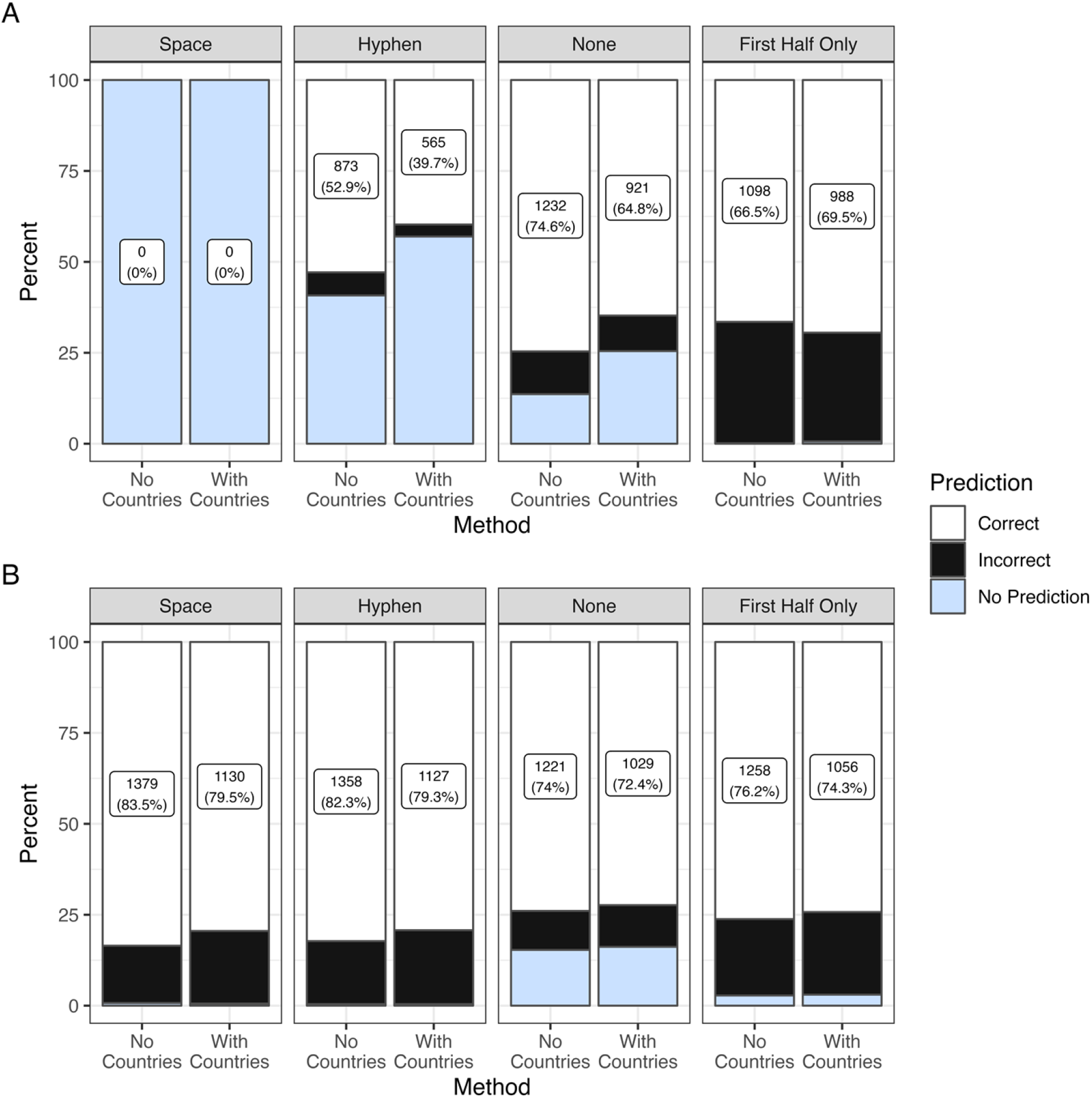
Accuracy of Gender Predictions Based on Delimiter between Two-Part Names. Panel A is Genderize and Panel B is Gender API. Plot facets correspond to the type of delimiter separating two-part names. Stacked bars correspond to correct, incorrect, and no predictions respectively. Bars are labeled with the count and percent of correct gender predictions.

The accuracy of Genderize and Gender API were evaluated for statistical significance by comparing the percent of correct gender predictions between delimiter categories. The difference in Gender prediction accuracy between Genderize and Gender API when evaluating two-part names without a delimiting character and including countries in the API call was not significant. All other comparisons between Genderize and Gender API were significant in favor of Gender API (p<0.001).

### Gender prediction accuracy by API-reported confidence thresholds

There was high agreement overall between gender prediction services and the gold standard labeled dataset (**Fig 3**). Genderize reported over 50% confidence in gender predictions for 32,573 (98.8%) trialists. Similarly, Gender API reported over 50% confidence for 32,587 (98.8%) trialists. Gender API demonstrated a correlation of 0.91 between its reported confidence and actual accuracy, compared to Genderize’s correlation of 0.82. The Brier scores for Gender API and Genderize were 0.0077 and 0.0048 respectively.

**Fig 3:**
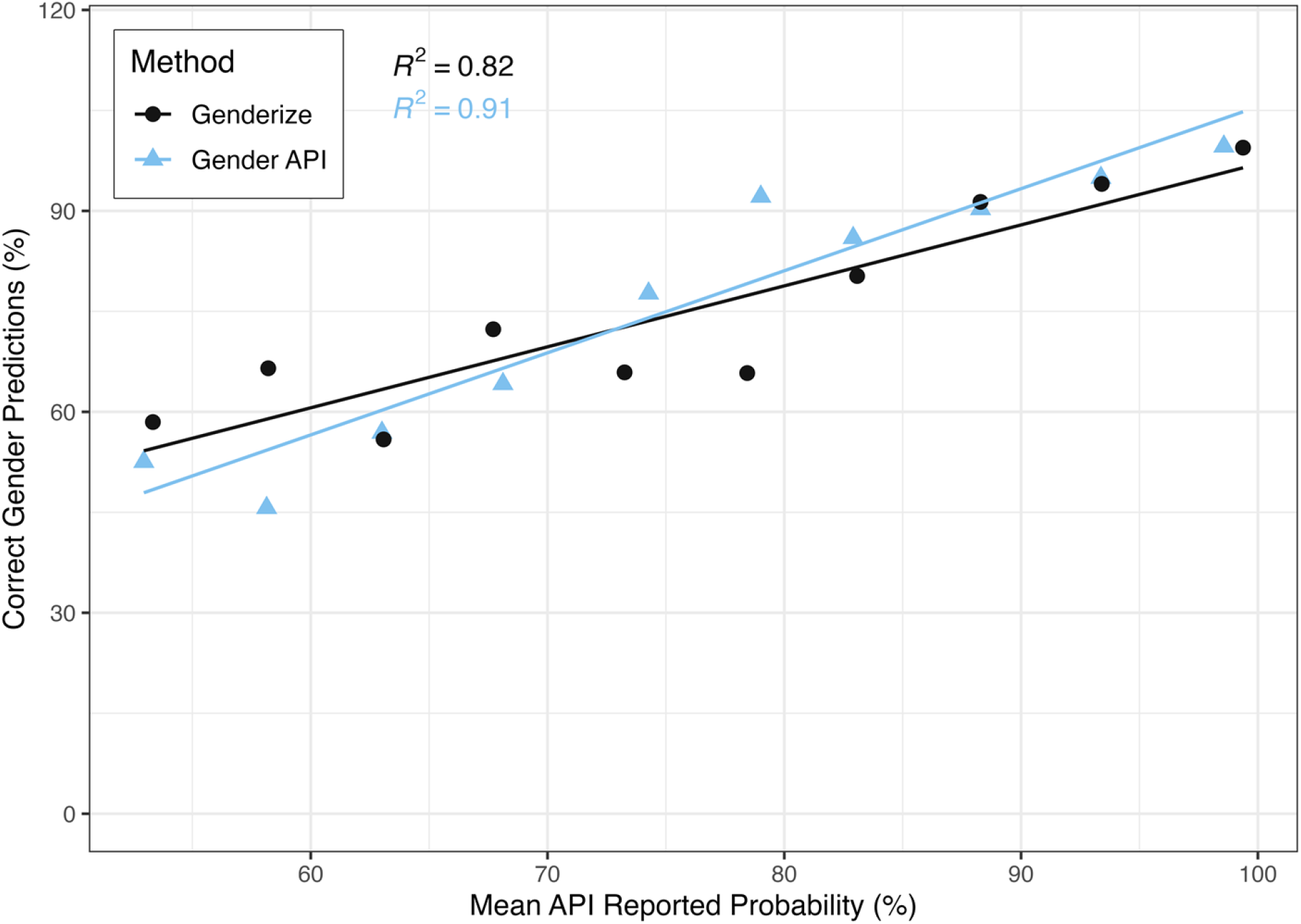
Experimental Name Prediction Accuracy At Different API Probability Cutoffs. Names with gender predictions were aggregated into the following API reported probability bins: 50%-55%, 55%-60%, 60%-65%, 65%-70%, 70%-75%, 75%-80%, 80%-85%, 85%-90%, 90%-95%, 95%-100%. The API reported probabilities within each bin were averaged and plotted on the x-axis. The experimentally determined gender prediction accuracies for the names in each bin are visualized on the y-axis.

### Replication of analyses by Santamaria 2018 and Sebo 2021

The original dataset used by Santamaria consisted of 5779 first names with known genders, 34% of whom were women. Only 0.4% of the 5779 had diacritical marks. In addition, 1.1% and 2% of names contain spaces or hyphens, respectively. The original paper reported 80% accuracy using Genderize and 87% using Gender API. In our re-analysis, Genderize predicted the correct gender 92.5% of the time. Similarly, Gender API achieved 92.8% accuracy. The difference in accuracy between Genderize and Gender API was not statistically significant.

The dataset originally analyzed by Sebo 2021 included 6131 names of whom 50.3% were women. Diacritical marks were present in 6.6% of names. 10.2% of names contained spaces, and 6.6% of names included a hyphen. The original paper reported 81% accuracy using Genderize and 97% with Gender API. In our re-analysis, the accuracy of Genderize and Gender API on these 6131 names were 86.2% and 98% respectively. McNemar’s test indicated that the differences in accuracy was statistically significant (p<0.001), in favor of Gender API. Gender API was 99.5% accurate when evaluating names with diacritical marks, while Genderize was 71.7% accurate.

### Cost and Accessibility

Genderize and Gender API provide a graphical user interface, while the gender R package requires programming. Genderize [10] provides 1,000 free predictions per day, whereas Gender API [11] only allows 100 free predictions per month. Gender API currently costs 4.85x more than Genderize for a monthly subscription that provides 100,000 predictions.

## Discussion

Genderize and Gender API both demonstrated over 95% overall accuracy on our gold-standard dataset of cancer clinical trialists. Genderize was slightly more accurate than Gender API when countries were not included in the API call. Conversely, Gender API performed slightly better than Gender API when countries were included. For both services, including countries reduced the number of incorrect gender assignments at the cost of increasing the number of names with no predicted gender (**Fig 1, Supplementary Table 2, Supplementary Table 3**). The gender R package performed worse than Genderize or Gender API (**Table 1**).

Genderize and Gender API differed in how their accuracy was affected by the delimiter separating two-part first names. Genderize was most accurate when two-part first names were appended together without a delimiter (**Fig 2**). In fact, Genderize appeared to be incompatible with two-part first names that were delimited by a space as the service yielded zero correct predictions when evaluating such names. Conversely, Gender API performed best when two-part first names were delimited with a space. The slightly higher overall gender prediction accuracy attained by Genderize compared to Gender API is partially an artifact of our decision to append two-part names without a delimiter in the baseline comparison, since Gender API performed best when two-part names were delimited with a space.

A commonality between this analysis and several previous studies was the lower prediction accuracy of Genderize and Gender API when evaluating Asian names, with the exception of Japanese names [15,19]. The higher accuracy achieved for both services in our re-analysis of Santamaria’s dataset indicates that both services have improved since 2015, although Genderize improved by a larger margin. Gender API outperformed Genderize when re-analyzing Sebo’s dataset largely because Gender API handled two-part names that were delimited by spaces as well as names with diacritical marks. In fact, a follow up study [14] by the same author recommended removing diacritical marks and modifying two-part names to improve the accuracy of Genderize.

This study’s results should be interpreted with certain caveats in mind. We did not filter out recurring first names during this analysis because the count of names in real-world datasets like ours tends to follow a long-tail distribution [29]. The process for determining the “gold-standard” gender of each of the trialists relied on inference from available information. Affiliation data was missing for a substantial subset of authors, mostly due to the older practice of MEDLINE including only the affiliation of the first author; a substantial number of high-profile oncology journals (e.g., the *Journal of Clinical Oncology* and *Blood*) did not include clear 1:1 mappings for author-to-affiliation for a period of time; this issue affects at least 320 (4%) of manuscripts in the HemOnc KB. A substantial subset of recently added authors to the HemOnc KB have not had their gender determined yet (13.7%), and this subset has some important differences from the set of determined genders. Most notably, the undetermined subset has many more Asian hyphenated names (43.8% vs 3.8%) and authors with a country of affiliation including South Korea, China, Singapore, and/or Taiwan (45.6% vs 3.42%). It is thus likely that our results represent a “best-case scenario” and that automated gender mapping will become increasingly difficult as cancer clinical trials are increasingly conducted in the Asia-Pacific region [30,31]. Additionally, a researcher’s nationality in our data set does not always reflect the cultural origin of their first name as some researchers immigrated to the country of their academic affiliation.

It is important to note that Genderize, Gender API, and the gender R package assume a gender binary. However, a recent survey [32] found that 1.6% of U.S. adults identify as transgender or nonbinary. With new algorithmic advancements such as Genderize and Gender API, it is imperative that inclusivity is incorporated. Going forward, tools that infer gender based on name should be trained on data that include trangender and nonbinary people, and they should include the option to predict an individual as non-binary or transgender. Gender prediction is not simply a binary classification problem. Transgender individuals will likely make up a small percentage of the dataset of names, which could make obtaining a correct prediction for these individuals very challenging. Yet, by not incorporating them, we are excluding countless people from this algorithm and ensuring the prediction of their gender to have a 0% accuracy.

Both Genderize and Gender API demonstrated high gender prediction accuracy with Western names that were highly normalized without middle or last names or diacritical marks. The cost per name evaluated with Genderize is also several times cheaper than Gender API. However, Genderize loses accuracy compared to Gender API when name formatting becomes less consistent. The SSA and IPUMS methods of the gender R package were less accurate but are open-source alternatives. The results from this study provide a new benchmark for gender inference tools.

## Data Availability

The data that support the findings of this study are publicly available from the Harvard DataVerse: https://dataverse.harvard.edu/privateurl.xhtml?token=6d620f82-5ef2-4ea6-90a5-19fb8ca4fe80

**Alexander D. VanHelene:** Conceptualization, Methodology, Software, Validation, Formal analysis, Investigation, Data Curation, Writing - Original Draft, Writing - Review & Editing, Visualization. **Ishaani Khatri:** Conceptualization, Writing - Original Draft, Writing - Review & Editing. **C. Beau Hilton:** Formal analysis, Writing - Review & Editing. **Sanjay Mishra:** Methodology, Formal analysis, Investigation, Data Curation, Visualization, Writing - Review & Editing, Funding acquisition. **Ece D. Gamsiz Uzun:** Visualization, Writing - Review & Editing. **Jeremy L. Warner:** Conceptualization, Methodology, Validation, Formal analysis, Investigation, Resources, Data Curation, Writing - Original Draft, Writing - Review & Editing, Visualization, Supervision, Funding acquisition.

## Acknowledgements

We would like to acknowledge the efforts of the editorial board of HemOnc.org.

